# Workplace measures against COVID-19 during the winter third wave in Japan: company size-based differences

**DOI:** 10.1101/2021.02.14.21251716

**Authors:** Tomohiro Ishimaru, Masako Nagata, Ayako Hino, Satoshi Yamashita, Seiichiro Tateishi, Mayumi Tsuji, Akira Ogami, Shinya Matsuda, Yoshihisa Fujino, for the CORoNaWork Project

## Abstract

**Objectives:** Little is known about workplace measures against coronavirus disease 2019 (COVID-19) in Japan after the first state of emergency period, especially in micro-, small-, and medium-sized enterprises (MSMEs). This study aimed to provide an overview of the current situation of anti-COVID-19 measures in Japanese enterprises, considering company size.

**Methods:** This study was an Internet-based nationwide cross-sectional study. Data were collected using an online self-administered questionnaire in December 2020 during the third wave of COVID-19. The chi-squared test for trend was performed to calculate the *p*-value for trend for each workplace measure across company sizes.

**Results:** For the 27,036 participants, across company sizes, the most prevalent workplace measure was encouraging mask wearing at work, followed by requesting that employees refrain from going to work when ill and restricting work-related social gatherings and entertainment. These measures were implemented by approximately 90% of large-scale enterprises and by more than 40% of micro- and small-scale enterprises. In contrast, encouraging remote working and restricting eating and drinking at personal workspaces were implemented by less than half of large-scale enterprises and by around 15% of micro- and small-scale enterprises. There were statistically significant differences in all workplace measures by company size (all *p*-values < .001).

**Conclusions:** We found that various responses to COVID-19 had been taken in workplaces. However, some measures, including remote working, were still not well implemented, especially in smaller enterprises. The findings suggest that occupational health support for MSMEs is urgently needed to mitigate the current wave of COVID-19.

## Introduction

Coronavirus disease 2019 (COVID-19) is an infectious disease caused by SARS-CoV-2 that is easily transmitted between persons; therefore, infection prevention and control in the workplace are of major concern.^1^ A previous study in Japan reported that most companies had taken individual-level precautions, such as hand washing and cough etiquette, but lagged behind in terms of organizational-level initiatives such as remote working and staggered commuting, especially in micro-, small-, and medium-sized enterprises (MSMEs), because of limited resources to respond to COVID-19.^2^ However, this previous study was conducted in mid-March 2020 during the early stages of the epidemic, and the current situation may differ.

A state of emergency was first declared in some areas of Japan on April 7, 2020; this declaration was later extended to the entire country until May 25, 2020.^3^ Although Japan did not impose a mandatory lockdown, many companies followed the voluntary-basis request from the government during the period, which included temporary closures and restrictions on business.^4^ One reason for the relatively low COVID-19 infection rate in Japan may be the corporate infection control efforts in the workplace.^5^ However, little is known about the current situation regarding workplace measures in Japan after the first state of emergency declaration, especially in MSMEs. Therefore, the purpose of this study was to provide an overview of the current situation of measures against COVID-19 in Japanese enterprises, taking company size into account. The findings will offer evidence on good practice for balancing business and infection control. The results will be useful for both countries and workplaces struggling during the pandemic.

## Subjects and Methods

### Study design and participants

This study was a part of the Collaborative Online Research on the Novel-coronavirus and Work (CORoNaWork) Project. Fujino et al. have introduced the details of the study protocol elsewhere.^6^ In brief, the CORoNaWork Project is an Internet-based nationwide prospective cohort study in Japan. The present cross-sectional study used data from the baseline survey, which was performed in December 2020. A total of 33,087 participants selected using cluster sampling with stratification by sex, age, region, and job type answered the online self-administered questionnaire. Panelists registered as health care workers or caregivers were not invited to participate in the survey. After excluding invalid responses, 27,036 participants were eligible for the analysis. When this survey was conducted, the numbers of COVID-19 infections and deaths were much higher than in the first and second waves; therefore, Japan was on maximum alert during the third wave.

### Questionnaire

This study used questionnaire data on sex, age, postal code of workplace, job type, company size, and workplace measures. Postal code was used to identify the geographical region of each workplace. The participants reported their job type as mainly desk work, work involving communicating with people, or manual work. Company size was classified as micro-scale (< 10 employees), small-scale (10–49 employees), medium-scale (50–999 employees), or large-scale (≥ 1,000 employees).

An original list of workplace measures was developed. We first prepared an initial list based on relevant publications listing standard workplace measures against COVID-19 in Japan.^2, 7, 8^ Subsequently, we developed the draft list in consultation with an expert panel on the basis of their practical experience. Finally, we selected 10 prioritized items, and all authors approved the final list. The question assessing workplace measures was as follows: “Are any of the following measures against COVID-19 currently taken at your workplace?” For each measure listed, the response options were *yes* or *no*.

### Data analysis

The participants’ demographic information is shown using counts and percentages. We compared the percentage for each of the 10 workplace measures against COVID-19 by company size. Chi-square tests for trend were performed to calculate the *p*-values for trend for each workplace measure across company sizes. Statistical significance was assessed at *p*-value < 0.05. Stata/SE 16.1 (StataCorp, College Station, TX, USA) was used for the statistical analysis.

## Results

Of the 27,036 participants, approximately half were men (51.1%) and around half engaged in desk work (49.8%) (Table 1). Regarding company size, 22.8% of the participants worked at micro-scale companies, 16.2% worked at small-scale companies, 35.9% worked at medium-scale companies, and 25.1% worked at large-scale companies.

**Table 1.** Demographic characteristics of the participants

|  | *n* | (%) |
| --- | --- | --- |
| Sex |  |  |
| Men | 13,814 | (51.1) |
| Women | 13,222 | (48.9) |
| Age (years) |  |  |
| 20–29 | 1,905 | (7.0) |
| 30–39 | 4,858 | (18.0) |
| 40–49 | 8,011 | (29.6) |
| 50–59 | 9,012 | (33.2) |
| 60–65 | 3,250 | (12.0) |
| Geographical region of workplace |  |  |
| Hokkaido | 695 | (2.6) |
| Tohoku | 2,346 | (8.7) |
| Kanto | 6,322 | (23.3) |
| Chubu | 4,894 | (18.1) |
| Kansai | 3,180 | (11.8) |
| Chugoku and Shikoku | 3,379 | (12.5) |
| Kyushu and Okinawa | 2,305 | (8.5) |
| Unknown | 3,915 | (14.5) |
| Job type |  |  |
| Desk work | 13,468 | (49.8) |
| Communication work | 6,927 | (25.6) |
| Manual work | 6,641 | (24.6) |
| Company size (number of employees) |  |  |
| 1–9 | 6,165 | (22.8) |
| 10–49 | 4,390 | (16.2) |
| 50–999 | 9,703 | (35.9) |
| ≥ 1000 | 6,778 | (25.1) |

Table 2 displays the number and percentage of respondents reporting each workplace measure against COVID-19 by company size. For all company sizes, the most prevalent workplace measure was encouraging mask wearing at work, followed by requesting that employees refrain from going to work when ill and restricting work-related social gatherings and entertainment. These measures were implemented by approximately 90% of large-scale enterprises and by more than 40% of micro-scale enterprises. In contrast, encouraging remote working and restricting eating and drinking at personal workspaces were implemented by less than half of large-scale enterprises and by around 15% of micro- and small-scale enterprises. There were statistically significant differences in all 10 workplace measures across companies of different sizes (all *p*-values < .001).

**Table 2.** Workplace measures against COVID-19 by company size

|  | Company size (number of employees) |  |  |  | *p*-value<br>for trend |
| --- | --- | --- | --- | --- | --- |
|  | 1–9<br>*n* = 6,165 | 10–49<br>*n* = 4,390 | 50–999<br>*n* = 9,703 | ≥ 1000<br>*n* = 6,778 |  |
| Encouraging mask wearing at work, *n* (%) | 3,319 (53.8) | 3,303 (75.2) | 8,452 (87.1) | 6,149 (90.7) | < .001 |
| Requesting that employees refrain from going to work<br>when ill, *n* (%) | 2,972 (48.2) | 3,052 (69.5) | 8,103 (83.5) | 6,103 (90.0) | < .001 |
| Restricting work-related social gatherings and<br>entertainment, *n* (%) | 2,743 (44.5) | 2,838 (64.6) | 7,709 (79.4) | 5,924 (87.4) | < .001 |
| Enforcing temperature measurement, *n* (%) | 2,068 (33.5) | 2,543 (57.9) | 7,308 (75.3) | 5,330 (78.6) | < .001 |
| Installing partitions or changing the working environment<br>(e.g., desk layout or flow lines), *n* (%) | 1,908 (30.9) | 2,059 (46.9) | 6,457 (66.5) | 5,310 (78.3) | < .001 |
| Restricting face-to-face meetings, *n* (%) | 1,835 (29.8) | 1,782 (40.6) | 6,036 (62.2) | 5,024 (74.1) | < .001 |
| Stopping business trips, *n* (%) | 1,757 (28.5) | 1,836 (41.8) | 6,056 (62.4) | 5,011 (73.9) | < .001 |
| Arranging health screenings for visitors, *n* (%) | 1,496 (24.3) | 1,498 (34.1) | 5,279 (54.4) | 4,026 (59.4) | < .001 |
| Encouraging remote working, *n* (%) | 1,314 (21.3) | 652 (14.9) | 2,556 (26.3) | 3,272 (48.3) | < .001 |
| Restricting eating and drinking at personal workspaces, *n*<br>(%) | 677 (11.0) | 511 (11.6) | 1,843 (19.0) | 1,566 (23.1) | < .001 |

## Discussion

The current study provides an overview of workplace measures against COVID-19 during the winter third wave in Japan, taking company size into account. We found that, especially in large-scale enterprises, various responses to COVID-19 had already been taken at workplaces, including encouraging mask wearing at work, requesting that employees refrain from going to work when ill, and restricting work-related social gatherings and entertainment. Our results are similar to those reported in a previous study on workplace measures conducted in mid-March 2020 (encouraging mask wearing at work: 80.2%; requesting that employees refrain from going to work when ill: 76.4%).^2^ These measures align with recommendations of the national campaign during the first state of emergency period, which included avoiding “the 3Cs” (closed spaces, crowded places, and close-contact settings).^3^ Thus, these types of workplace measures have been implemented since the early stage of the epidemic.

Another remarkable finding of this study is that smaller enterprises were less likely to have implemented workplace measures against COVID-19. The finding is consistent with a previous study conducted in mid-March 2020.^2^ This finding therefore indicates that MSMEs did not make much progress in terms of measures against COVID-19 since the first state of emergency declaration. One possible reason for this lack of progress is that MSMEs often face difficulty in implementing occupational health activities because of a lack of financial, human capital, and technological resources.^9^ In Japan, the requirements for occupational health staff members depend on the number of employees in a workplace: ≥ 1,000 employees—a full-time occupational physician, 50–999 employees—a part-time occupational physician, ≥ 50 employees—a health officer, and 10–49 employees—a health promoter.^10^ Therefore, the results of the present study may be attributed to the difference in occupational health staff members by company size. This finding suggests that occupational health support from external resources is urgently needed for MSMEs. For example, the development of simple tools for infection prevention, mechanisms for external occupational health experts to offer advice, and financial support for workplace measures can be considered.

Some measures, such as remote working and restrictions on eating and drinking at personal workspaces, had still not been implemented by the majority of companies—even large-scale enterprises. Few studies have investigated the proportion of companies imposing restrictions on eating and drinking at personal workspaces, but the implementation of remote working has not changed much, compared with the results of previous studies conducted around the first state of emergency period.^2, 4^ The finding suggests potential obstacles to promoting remote working, such as cultural barriers and administrative difficulties.^11^ Cultural barriers may exist for both employers and employees.

For example, employers may excessively demand that work to be carried out on site, and workers are willing to work at the office.^12^ Interestingly, the current study found that the frequency of encouraging remote work was lower in workplaces with 10–49 employees (14.9%) than in workplaces with nine or fewer employees (21.3%). We considered the possibility that administrative difficulties might be more likely to occur in small-scale enterprises than in micro-scale enterprises. However, remote working is effective not only during a pandemic but also during large earthquakes and other disasters.^13^ In terms of business continuity, future research is warranted on the further expansion of remote working, especially in small-scale enterprises.

This study has several limitations. First, the current study did not use random sampling or collect data from all companies. Consequently, the sample may not represent the national situation, and any generalization of the results should be carried out with care. For example, there is a risk of overestimation if multiple study participants were from the same organization. To cope with these problems, the current study was conducted using cluster sampling with stratification by sex, age, region, and job type. Second, the current study did not evaluate all types of workplace measures; for instance, information dissemination and actions for confirmed COVID-19 cases were not considered. However, because we focused on preventive measures listed in guidelines and relevant publications in Japan,^2, 7, 8^ we believe that these items reflect the current situation of measures against COVID-19 at each company.

In conclusion, this study revealed the current situation regarding workplace measures against COVID-19 in Japan. We found that various responses to COVID-19 have been implemented at workplaces. However, some measures, including remote working, were still not well implemented, especially in relatively small enterprises. The findings suggest that occupational health support for MSMEs is urgently needed to mitigate the current wave of COVID-19.

## Data Availability

No additional data are available.

## Acknowledgments

This study was supported by a research fund from the University of Occupational and Environmental Health, Japan; General Incorporated Foundation (Anshin Zaidan): The Development of Educational Materials on Mental Health Measures for Managers at Small-sized Enterprises; Health, Labour and Welfare Sciences Research Grants: Comprehensive Research for Women’s Healthcare (H30-josei-ippan-002); Research for the Establishment of an Occupational Health System in Times of Disaster (H30-roudou-ippan-007), and scholarship donations from CHUGAI PHARMACEUTICAL CO., LTD.

The current members of the CORoNaWork Project, in alphabetical order, are as follows: Dr. Yoshihisa Fujino (present chairperson of the study group), Dr. Akira Ogami, Dr. Arisa Harada, Dr. Ayako Hino, Dr. Chimed-Ochir Odgerel, Dr. Hajime Ando, Dr. Hisashi Eguchi, Dr. Kazunori Ikegami, Dr. Keiji Muramatsu, Dr. Koji Mori, Dr. Kyoko Kitagawa, Dr. Masako Nagata, Dr. Mayumi Tsuji, Dr. Rie Tanaka, Dr. Ryutaro Matsugaki, Dr. Seiishiro Tateishi, Dr. Shinya Matsuda, Dr. Tomohiro Ishimaru, Dr. Tomohisa Nagata, Dr. Yosuke Mafune, and Ms. Ning Liu. All members are affiliated with the University of Occupational and Environmental Health, Japan.

## Disclosure

### Approval of the research protocol

This study was approved by the Ethics Committee of the University of Occupational and Environmental Health, Japan. *Informed Consent*: Informed consent was obtained from all participants. *Registry and the Registration No. of the study/trial*: N/A. *Animal Studies*: N/A. *Conflict of Interest*: N/A.

## Notes

### Competing Interest Statement

The authors have declared no competing interest.

### Author Declarations

This study was approved by the Ethics Committee of the University of Occupational and Environmental Health, Japan.

## References

1. Wu D, Wu T, Liu Q, Yang Z. The SARS-CoV-2 outbreak: What we know. Int J Infect Dis 2020;94:44–48.

2. Sasaki N, Kuroda R, Tsuno K, Kawakami N. Workplace responses to COVID-19 and their association with company size and industry in an early stage of the epidemic in Japan. Environmental and Occupational Health Practice 2020;2:peohp.2020-0007-OA.

3. Shimizu K, Negita M. Lessons Learned from Japan’s Response to the First Wave of COVID-19: A Content Analysis. Healthcare (Basel) 2020;8.

4. Nomura S, Yoneoka D, Tanoue Y, et al. Time to Reconsider Diverse Ways of Working in Japan to Promote Social Distancing Measures against the COVID-19. J Urban Health 2020;97:457–460.

5. World Health Organization. WHO Coronavirus Disease (COVID-19) Dashboard. https://covid19.who.int/. xPublished January 2020. Accessed February 1, 2021.

6. Fujino Y, Ishimaru T, Eguchi H, et al. Protocol for a nationwide Internet-based health survey in workers during the COVID-19 pandemic in 2020. medRxiv 2021:2021.02.02.21249309.

7. Japan Society for Occupational Health, Japanese Society of Travel Medicine. Information of new coronavirus infection. https://www.sanei.or.jp/. xPublished December 2020. Accessed February 1, 2021.

8. Ministry of Health Labour and Welfare, Japan. Checklist for the prevention of COVID-19 spreading at workplaces. https://www.mhlw.go.jp/content/000694987.pdf. Published November 2020. Accessed February 1, 2021.

9. Hasle P, Limborg HJ. A review of the literature on preventive occupational health and safety activities in small enterprises. Ind Health 2006;44:6–12.

10. Muto T. Status and Future Challenges of Japanese Occupational Health Services. Policy and Practice in Health and Safety 2007;5:169–180.

11. Chung H, van der Horst M. Women’s employment patterns after childbirth and the perceived access to and use of flexitime and teleworking. Human relations; studies towards the integration of the social sciences 2018;71:47–72.

12. Lott Y, Abendroth A-K. The non-use of telework in an ideal worker culture: why women perceive more cultural barriers. Community, Work & Family 2020;23:593–611.

13. Anan T, Mori K, Kajiki S, Tateishi S. Emerging Occupational Health Needs at a Semiconductor Factory Following the 2016 Kumamoto Earthquakes: Evaluation of Effectiveness and Necessary Improvements of List of Postdisaster Occupational Health Needs. J Occup Environ Med 2018;60:198–203.

